# Prevalence of severe adverse events in health professionals after receiving the first dose of the COVID-19 vaccination in Togo, March 2021

**DOI:** 10.1101/2021.04.20.21254863

**Authors:** Yao Rodion Konu, Fifonsi Adjidossi Gbeasor-Komlanvi, Mouhoudine Yerima, Arnold Sadio, Martin Kouame Tchankoni, Wendpouire Ida Carine Zida-Compaore, Josée Nayo-Apetsianyi, Kossivi Agbélénbko Afanvi, Sibabe Agoro, Mounerou Salou, Dadja Essoya Landoh, Atany B. Nyansa, Essohanam Boko, Moustafa Mijiyawa, Didier Koumavi Ekouevi

## Abstract

**Introduction:** Covid-19 vaccines can cause adverse events (AE) that can lead to increased hesitation or fear of vaccination. This study aims at estimating the prevalence of severe adverse events (SAEs) and their associated factors among health professionals (HPs) vaccinated with COVISHIELD™ vaccine in Togo.

**Methods:** A cross-sectional study was conducted from March 13^th^ to 19^th^, 2021 in Togo among HPs who received the first dose of vaccine. An online self-administered questionnaire was used to collect data on sociodemographic characteristics and vaccination. SAEs were defined as one resulting in hospitalization, medical consultation, or inability to work the day following the administration of the vaccine. Regression analysis were performed to assess factors associated with SAEs.

**Results:** A total of 1,639 HP (70.2% male) with a median age [IQR] of 32 years [27-40] participated. At least one AE was reported among 71.6% (95%CI= [69.3-73.8]). The most commonly reported AEs were pain at the injection site (91.0%), asthenia (74.3%), headache (68.7%), soreness (55.0%), and fever (47.5%). An increased libido was also reported in 3.0% of HP. Among HP who experienced AEs, 18.2% were unable to go to work the day after vaccination, 10.5% consulted a medical doctor, and 1.0% were hospitalized. The SAE prevalence was 23.8% (95%CI= [21.8-25.9]). Being <30 years (aOR=5.54; p<0.001), or 30-49 years (aOR=3.62; p<0.001) and being female (aOR=1.97; p<0.001) were associated with SAEs.

**Conclusion:** Despite the occurrence of SAEs, current data collected in Togo about adverse events are reassuring with COVISHIELD™ vaccine and how they could be managed.

**Key points:**

- The prevalence of severe adverse events (SAEs) after COVID-19 vaccination was high among health professionals in Togo, especially in the youths (under 30years).
- Despite the occurrence of SAEs, current data collected in Togo about adverse events are reassuring with COVISHIELD(TM) vaccine and provide information about how the could be managed.

## 1. Introduction

Coronavirus disease 2019 (COVID-19) was first described in China in December 2019 [1] and resulted in the declaration of a pandemic in January 2020 [2]. As of March 24, 2021, more than 120 million cases of COVID-19 infections, with more than 2 million deaths, were reported worldwide [3].

No cure or vaccine was available until December 2020. Therefore, controlling the infection to prevent the spread of COVID-19 was considered the only intervention [4]. Several social and public health risk mitigation measures were proposed and implemented to reduce the spread of the pandemic. These measures include individual measures (frequent hand hygiene, physical distancing, and use of masks) and physical and social distancing measures (reduction of mass gatherings, and promotion of telecommuting) [5].

However, given that vaccination has been identified as a relevant intervention for stopping epidemics and fighting against infectious diseases, research on a COVID-19 vaccine has been promoted. At an early stage of this pandemic, the World Health Organization (WHO) and the Global Alliance for Vaccines and Immunization (Gavi) launched a pooled procurement mechanism for new COVID-19 vaccines called the COVAX Facility to ensure fair, global and equitable access to vaccines [6]. This facility also aims to end the acute phase of the COVID-19 pandemic by accelerating the development of safe and effective vaccines against COVID-19 and contributing to the development of production capacity [7]. The first vaccines received the Emergency Use Listing from WHO in December 2020 [8].

Like many African countries, Togo has joined the COVAX facility and participated in COVID-19 vaccine procurement process [6]. On March 7, 2021, Togo received the first allocation of 156,000 doses of the SII-AstraZeneca (COVISHIELD™) vaccine [9]. COVISHIELD™ was developed by AstraZeneca and Oxford University and produced by the Pune-based Serum Institute of India [10]. It is a viral vector vaccine (ChAdOx1-S) that was approved for emergency use by the WHO [11]. In Togo, the vaccination campaign was launched on March 10^th^, 2021. According to the National Deployment and Vaccination Plan for COVID-19 vaccines, the priority target was 35,119 health professionals (HP) followed by people over 50 years of age and those living with at least one comorbidity condition (hypertension, diabetes, heart disease etc.) [12]. These groups were targeted because they are at greater risk of exposure to COVID-19 or because they have an increased risk of death in case of infection.

Vaccination against COVID-19 is an essential pillar for controlling the pandemic in addition to other infection risk mitigation measures included in test, treat, track (T3) approach. In Togo, vaccination against COVID-19 is implemented at a time when the country is experiencing an epidemic peak with more than 600 cases per week since February 1^st^, 2021 compared to 100 cases per week in December 2020 [13]. As of March 24^th^, 2021, 9147 COVID-19 infections cases, of which, 105 deaths, have been reported in Togo [14].

Despite the awareness campaign on the importance of vaccination, several factors contribute to limiting the adherence of the population to this intervention. These include: i) the relative speed of discovery and availability the of vaccines, ii) the use of new technologies never before deployed in humans, and iii) the lack of hindsight regarding the safety of vaccines that are available.

As a result, false information has been circulating about these new vaccines and has contributed to the growing anxiety and vaccine hesitancy associated with a fear of occurrence of long-term adverse events. In addition, the suspension of the AstraZeneca vaccine in Europe after the occurrence of thrombosis cases has amplified psychosis with popular pressure to stop the vaccination campaign in Africa including Togo.

On March 11 and 12, Togo conducted a large-scale vaccination program against COVID-19 for HP and 18,249 were vaccinated [15]. In parallel with the surveillance of adverse events following immunization which is ensured by the pharmacovigilance service of the Ministry of Health, this study was launched to quickly document adverse events to reassure the population. Our objective was to estimate the prevalence of severe adverse events (SAEs) and their associated factors among HPs in Togo.

## 2. Methods

### 2.1. Study design and setting

This study was a cross-sectional study that was conducted from March 13 to 19, 2021 in Togo.

Togo is a country of West Africa that covers an area of 56,800 km^2^ with an average density of 145 inhabitants per square kilometer [16]. The population was 8.08 million in 2019, of which 50.2% were women [17]. Most of the population is young (60% of Togolese are under 25 years of age), and lives in rural areas (62%) [17]. Togo’s health system has a three-level pyramid structure: central, intermediate and peripheral levels. Administrative and healthcare delivery components are associated with every level.

### 2.2. Study population and sample size

The target population included all HPs who received the first dose of the COVISHIELD™ vaccine during the first phase of the vaccination campaign in Togo. For the purpose of this study, a health professional was defined as a professional who works or is affiliated with the health sector, including workers from health administration and logistics, clinical settings, and community health workers. The inclusion criteria for eligible participants were: (i) being a health professional aged 18 years and older; and (ii) having received the first dose of vaccine.

The sample size of participants was calculated using a single proportion population formula with a 95% confidence level. In the absence of previous studies on the severity of the adverse events of the vaccine against COVID-19, we hypothesized that 10% of the participants would experience at least one severe adverse event with a 2% margin error and a 10% nonresponse rate. The minimum number of participants was estimated at 960.

### 2.3. Data collection

A multiple-choice questionnaire was developed using items from a questionnaire of the Pharmacovigilance Department of the Togo Ministry of Health and from previous related studies found in the literature [18,19]. The questionnaire consisted of two main sections on sociodemographic and professional characteristics (age, sex, marital status, working place) and vaccination (date, process, confidence in the vaccine efficacy, occurrence of adverse events and likelihood of getting the second dose). The self-administered questionnaire was made available using a free online platform through the internal communication networks of the Ministry of Health. The data collection process began two days after the end of the first phase of the vaccine campaign which targeted HPs in Togo.

### 2.4. Definition of the outcome variable

The main outcome was the severity of reported adverse events. Adverse events were grouped into three categories: severe (presence of adverse events resulting in self-reported hospitalization, seeking medical consultation, and inability to work the day following the administration of the vaccine), moderate (presence of adverse events without hospitalization, consultation or impact on working ability), and no side effects.

### 2.5. Data management and statistical analysis

Data were imported into a Microsoft Excel database for data cleaning. Descriptive statistics were performed, and the results are presented using frequency tabulations and percentages for categorical variables. Quantitative variables are presented as medians with their interquartile range (IQR). Proportions were compared using Chi-square test or Fisher’s exact test when appropriate. Prevalence of SAEs was estimated with its 95% confidence interval (95% CI).

Univariable and multivariable logistic model regression were performed to assess factors associated with SAEs. For model building, characteristics that had a p-value <0.20 in univariable analysis were considered for the full multivariable models, which were subsequently finalized using a stepwise, backward elimination approach (p-value <0.05). Predictor variables were selected as those found to be relevant according to the literature review. Sensitivity analysis was performed based on a second definition of SAEs which considered only self-reported hospitalization or seeking medical consultation. Data analysis were performed using R^©^ version 4.0.1 software, and the level of significance was set at 5%.

### 2.6. Ethical considerations

Ethical approval was obtained from the ‘Comité de Bioéthique de Recherche en Santé’ (Bioethics Committee for Health Research) from the Togo Ministry of Health (No. 01/2021/CBRS). An introductory question was asked to ensure participants’ consent.

## 3. Results

### 3.1. Sociodemographic characteristics of health professionals

A total of 1,639 HPs (70.2% male) who had received the first dose of the vaccine responded to the questionnaire. This sample represents 4.6% of the HPs vaccinated in Togo.

The median age [IQR] of the participants was 32 years [27-40]. Approximately half (45.8%) of the participants resided or practiced in the *Grand-Lomé* region and 54.7% were married (Table 1).

**Table 1.** Sociodemographic characteristics of health professionals vaccinated against COVID-19 in Togo, 2021 (n = 1,639)

|  | Frequency | Proportion (%) |
| --- | --- | --- |
| **Age (years), Median (IQR)** | 32 (27-40) |  |
| **Age (years)** |  |  |
| <30 | 634 | 38.7 |
| 30-49 | 862 | 52.6 |
| ≥50 | 143 | 8.7 |
| **Sex** |  |  |
| Female | 488 | 29.8 |
| Male | 1151 | 70.2 |
| **Health region** |  |  |
| Grand-Lomé | 751 | 45.8 |
| Maritime | 243 | 14.8 |
| Plateaux | 161 | 9.8 |
| Centrale | 104 | 6.4 |
| Kara | 274 | 16.7 |
| Savanes | 106 | 6.5 |
| **Marital status** |  |  |
| Single/Widowed/Divorced | 742 | 45.3 |
| Married | 897 | 54.7 |

IQR: interquartile range

### 3.2. Prevalence of adverse events

Among the 1,639 participants, 1174 (71.6%, 95% CI = [69.3-73.8]) reported at least one side effect. The most commonly reported adverse events were pain at the injection site (91.0%), asthenia (74.3%), headache (68.7%), body aches (55.0%), and fever (47.5%). An increase in libido was also reported in 3.0% of the participants (Figure 1). Of the participants who experienced adverse events, 18.2% were unable to go to work the day after vaccination, 10.5% consulted a medical doctor, and 1.0% were hospitalized. Thus, the prevalence of SAEs was 23.8% (95% CI = [21.8-25.9]).

**Figure 1.**
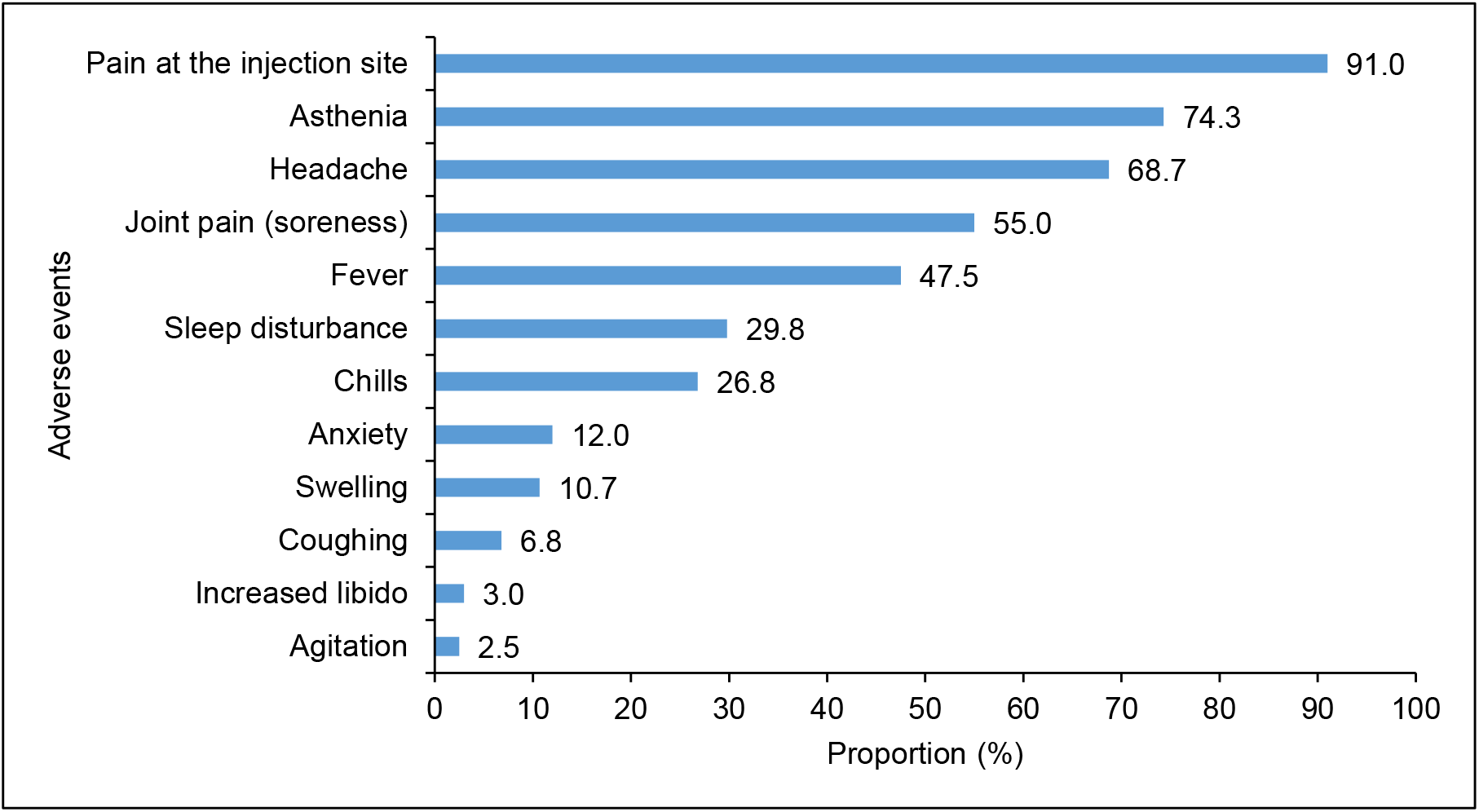
Proportion of adverse events among health professionals after vaccination against COVID-19, Togo, 2021 (n = 1,174)

### 3.3. Post vaccination treatment

Overall, medication use after vaccination was higher among participants with SAEs (p<0.001). The use of analgesics by participants with SAE was 62.3% vs 39.0% for participants with moderate events and 5.2% for those reporting no events (Table 2).

**Table 2.** Use of medication after vaccination against COVID-19 according to the severity of adverse events among health professionals, Togo, 2021

|  | Adverse events after COVID-19 vaccination |  |  |  | P |
| --- | --- | --- | --- | --- | --- |
|  | None | Moderate | Severe | Overall |  |
|  | (n = 442) | (n = 807) | (n = 390) | (n = 1,639) |  |
| Analgesics, n (%) | 23 (5.2) | 315 (39.0) | 243 (62.3) | 581 (35.4) | <0.001* |
| Antipyretics, n (%) | 15 (3.4) | 207 (25.7) | 181 (46.4) | 403 (24.6) | <0.001* |
| NSAIDs<sup>§</sup>, n (%) | 10 (2.3) | 82 (10.2) | 65 (16.7) | 157 (9.6) | <0.001* |
| Antihistamines, n (%) | 1 (0.2) | 23 (2.9) | 43 (11.0) | 67 (4.1) | <0.001** |
| Vitamin C, n (%) | 1 (0.2) | 19 (2.4) | 17 (4.4) | 37 (2.3) | <0.001** |
| Herbal medicines/infusions, n (%) | 0 (0.0) | 12 (1.5) | 14 (3.6) | 26 (1.6) | <0.001** |

<sup>§</sup>NSAIDs: non-steroidal anti-inflammatory drugs
\*Chi-squared test; \*\*Fisher test

### 3.4. Perception after vaccination

A total of 67.5% of survey participants expressed concern about long-term adverse events, and this concern was more pronounced among women than men (74.2% vs. 64.7%, p<0.001). Approximately one out of ten participants (10.9%) said they were not ready to take the second dose of the vaccine (Table 3).

**Table 3.** Perceptions among health professionals after vaccination against COVID-19, Togo, 2021

|  | **Female** | **Male** | **Overall** | **p** |
| --- | --- | --- | --- | --- |
|  | **(n = 488)** | **(n = 1,151)** | **(n = 1,639)** |  |
| **Afraid of long-term adverse events** |  |  |  | <0.001* |
| No | 126 (25.8) | 406 (35.3) | 532 (32.5) |  |
| Yes | 362 (74.2) | 745 (64.7) | 1107 (67.5) |  |
| **Regret to have been vaccinated** |  |  |  | <0.001* |
| No | 347 (71.1) | 923 (80.2) | 1270 (77.5) |  |
| Yes | 141 (28.9) | 228 (19.8) | 369 (22.5) |  |
| **Ready for the second dose** |  |  |  | 0.004* |
| Hesitation | 201 (41.2) | 391 (34.0) | 592 (36.1) |  |
| No | 59 (12.1) | 119 (10.3) | 178 (10.9) |  |
| Yes | 228 (46.7) | 641 (55.7) | 869 (53.0) |  |

\*Chi-squared test

### 3.5. Factors associated with the occurrence of severe adverse events

In univariate analysis, factors associated with the occurrence of SAEs included being under 30 years of age (OR=4.74; p<0.001), or 30-49 years of age (OR=3.00; p<0.001) and being female (OR=2.07; p<0.001). The same factors were associated with the occurrence of SAEs in multivariate analysis (Table 4).

**Table 4.**
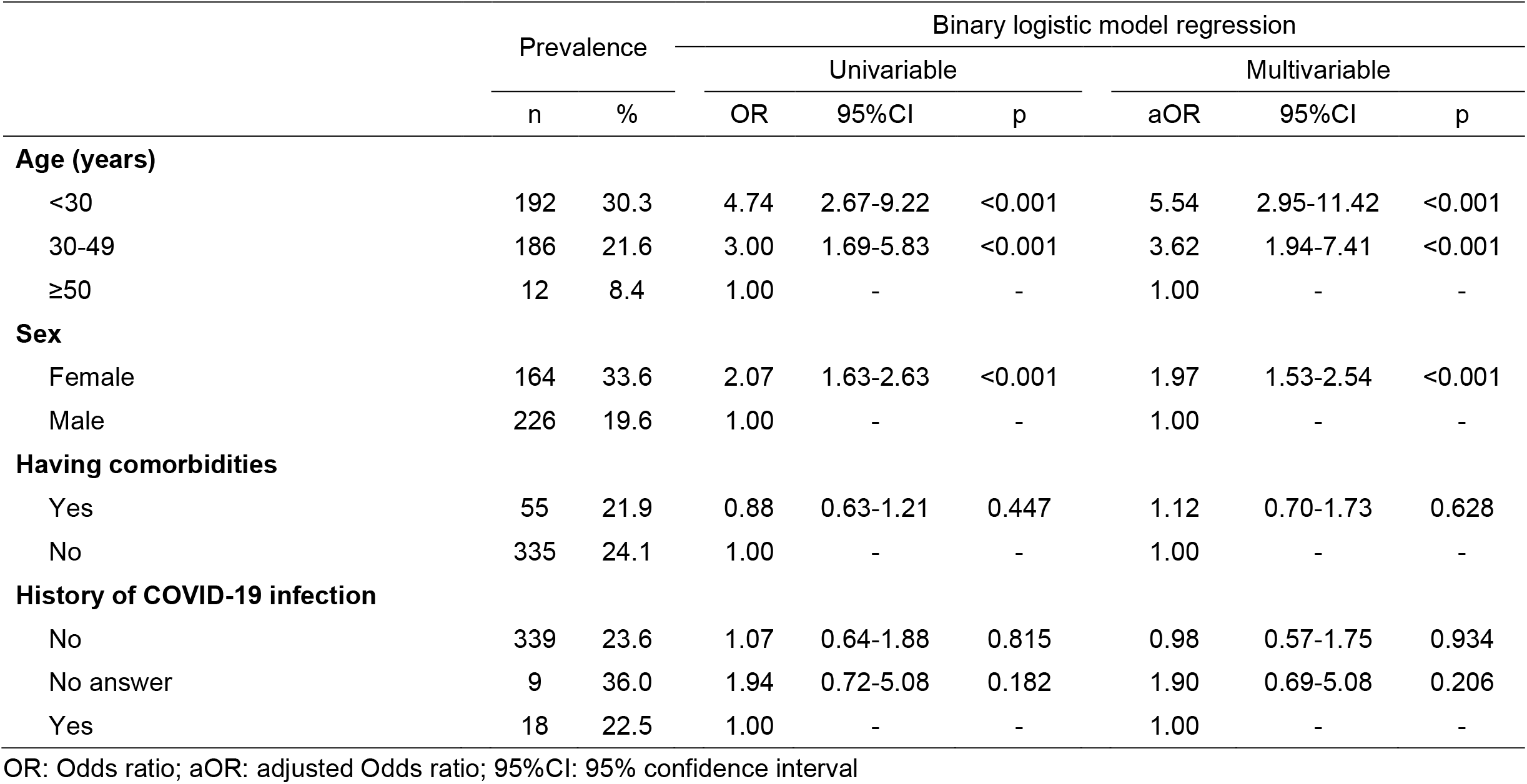
Factors associated with severe adverse events after COVID-19 vaccination among health professionals in Togo, 2021

Using the second definition of SAEs, the prevalence of SAEs was estimated at 10.6% (95% CI = [9.2-12.2]); the factors associated with the occurrence of SAE were the same both in univariate and in multivariate analysis (under 30 years of age aOR=2.23; p=0.049; 30-49 years of age, aOR=2.73, p=0.016 and being female, aOR=2.01, p<0.001).

## 4. Discussion

This is the first study in Africa to report SAEs after vaccination with the COVISHIELD™ vaccine in the context of COVID-19 among HPs. We included 4.6% of healthcare professionals vaccinated in Togo. SAEs were reported among 23.8% of participants in the current study. Most adverse events reported were mild or severe and compared with those reported in trials of AstraZeneca [20,21]. However, this study clearly shows that the odds of experiencing SAEs after vaccination against COVID-19 are higher in younger people and women compared to older adults and men, respectively.

Based on our operational definition, approximately one-quarter of participants reported SAEs. This value could be overestimated since because we included the inability to work the day after vaccination. If we consider only hospitalization and consultation of a medical doctor after vaccination, approximately 12.0% presented SAEs. In a single-blind, randomized, controlled, phase 2/3 trial (COV002), in the United Kingdom (UK), as of October 26, 2020, 13 serious adverse events occurred during the study period, but none of those were considered to be related to the study vaccine [22]. The interim analysis of the efficacy and safety of the ChAdOx1 nCoV-19 vaccine includes data from four ongoing blinded, randomized, controlled trials performed across three countries including the UK, Brazil and South Africa, and was reported in January 2021 [20]. In this analysis, 175 SAEs occurred in 168 participants, of which three events were classified as possibly related to a vaccine: one in the ChAdOx1 nCoV-19 group, one in the control group, and one in a participant who remained masked to group allocation.

Several definitions exist to classify adverse events according to their severity. For example, in the COV002 trial, in the UK, SAEs were defined as substantial limitations in activity and medical intervention or the requirement of therapy [22].

We did not include, the use of medication in our definition, given that the study population was included HPs who have easy access to medication and a proven tendency to self-medicate [23,24]. Interpretations of the severity of adverse events must take into account the definition we adopted.

In our study, SAEs were more common in younger people than in people aged 50 years and older. The younger the subject was, the more severe the adverse events tended to be. This trend has been reported by Oxford COVID Vaccine Trial Group and seems to be related to an exaggerated immune response in young subjects [22]. In other countries, severe events following the COVISHIELD™ vaccine have been observed more in subjects under 55 years of age, which has prompted France among other countries to exclusively recommend this vaccine for subjects aged 55 years and older. In Togo, such recommendation is difficult to apply given there is no other vaccine available to date.

Adverse events after vaccination were more pronounced in women. This observation is not unusual for vaccines in general [25] and seems to be consistent with data in the literature about COVID-19 vaccines [26,27]. More SAEs in women have been reported in other settings and appear to be related to a stronger immune response triggered by estrogen [25].

Most reported adverse events with the COVID-19 vaccine were mild. A sore arm was the most common, and others included headache, tiredness, and a mild flu-like symptoms [28,29]. Similar adverse events were reported in our study. However, in our study, an increase in libido was unexpectedly reported by 3.0% of HPs in both men and women. Among all UK spontaneous reports received between March 4,2021 and March 14,2021 for COVID-19 Oxford University/AstraZeneca vaccine, one case of increased libido, one of decreased libido and six cases of loss of libido were mentioned [30]. This information should be investigated to demonstrate an association with vaccination.

Concerning the perception/intention after the first dose of vaccine, two thirds of the participants expressed concern about long-term adverse events, and one out of ten declared they were not ready to take the second dose of the vaccine. These results could be explained by several concerns about ChAdOx1 nCoV-19 which arose in Europe and some African countries and resulted in the temporarily suspension the administration of this vaccine [31].

This study presents some limitations. We cannot exclude selection bias due to our sampling method. Another limitation is that data collection was based on a declarative approach. However, given that the participants were HPs, the reported effects are probably consistent with reality.

Finally, we used an operational definition to define SAE. This is a composite variable including absence from work the day after vaccination, medical consultation and hospitalization due to adverse events. This definition probably led to an overestimation of SAE, but this operational definition seems appropriate in the Togolese context.

Based on these results of a high prevalence of SAEs in young people, systematic prescription of antalgics or antipyretics could be proposed to young people who are eligible to be vaccinated. Careful monitoring of SAEs should be performed in younger people under 50 years of age. If another vaccine becomes available in Togo, the COVISHIELD™ vaccine should be reserved for older adults.

A cohort study is therefore needed for a follow-up of HPs to better document the occurrence of long-term adverse events. The results from the present study are useful for designing a sensitization program in order to reassure the general population about COVID-19 vaccination. At this stage, there is no reported sign in link with thrombosis.

## 5. Conclusion

In conclusion, despite the occurrence of SAEs, vaccination against COVID-19 remains an important strategy to fight against this pandemic. Current data collected in Togo are reassuring about adverse events and how they could be managed.

## Data Availability

Data are available on request to the corresponding author.

## Declarations

### i. Funding

Not applicable

### ii. Conflicts of interest/Competing interests

The authors declare that they have no competing interests. DEL works for the World Health Organization, Togo country office. The authors are solely responsible for the views expressed in this manuscript and do not necessarily represent the decisions, policies or views of the World Health Organization.

### iii. Availability of data and material

Available on request to the corresponding author.

### iv. Code availability

Not applicable

### v. Ethics approval

This study obtained ethical approval from the ‘Comité de Bioéthique de Recherche en Santé’ (Bioethics Committee for Health Research) from the Togo Ministry of Health (No. 01/2021/CBRS).

### vi. Consent to participate

informed consent was collected prior to inclusion.

### vii. Consent for publication

Not applicable

## Notes

### Competing Interest Statement

The authors have declared no competing interest.

### Funding Statement

No external funding was received.

### Author Declarations

Ethical approval was obtained from the Bioethics Committee for Health Research from the Togo Ministry of Health (No. 01/2021/CBRS).

## References

1. Zhu N, Zhang D, Wang W, Li X, Yang B, Song J, et al. A Novel Coronavirus from Patients with Pneumonia in China, 2019. N Engl J Med. 2020 Feb 20;382(8):727–33.

2. World Health Organization (WHO). Statement on the second meeting of the International Health Regulations (2005) Emergency Committee regarding the outbreak of novel coronavirus (2019-nCoV) [Internet]. [cited 2021 Mar 29]. Available from: https://www.who.int/news/item/30-01-2020-statement-on-the-second-meeting-of-the-international-health-regulations-(2005)-emergency-committee-regarding-the-outbreak-of-novel-coronavirus-(2019-ncov).

3. World Health Organization (WHO). WHO Coronavirus (COVID-19) Dashboard [Internet]. [cited 2021 Mar 24]. Available from: https://covid19.who.int

4. Bruinen de Bruin Y, Lequarre A-S, McCourt J, Clevestig P, Pigazzani F, Zare Jeddi M, et al. Initial impacts of global risk mitigation measures taken during the combatting of the COVID-19 pandemic. Saf Sci. 2020 Aug;128:104773.

5. World Health Organization (WHO). Overview of Public Health and Social Measures in the context of COVID-19 [Internet]. [cited 2020 Jun 19]. Available from: https://www.who.int/publications-detail-redirect/overview-of-public-health-and-social-measures-in-the-context-of-covid-19.

6. 172 countries and multiple candidate vaccines engaged in COVID-19 vaccine Global Access Facility [Internet]. [cited 2021 Mar 24]. Available from: https://www.who.int/news/item/24-08-2020-172-countries-and-multiple-candidate-vaccines-engaged-in-covid-19-vaccine-global-access-facility

7. World Health Organization (WHO). COVID-19 vaccines [Internet]. [cited 2021 Mar 24]. Available from: https://www.who.int/emergencies/diseases/novel-coronavirus-2019/covid-19-vaccines

8. World Health Organization (WHO). WHO issues its first emergency use validation for a COVID-19 vaccine and emphasizes need for equitable global access [Internet]. [cited 2021 Mar 26]. Available from: https://www.who.int/news/item/31-12-2020-who-issues-its-first-emergency-use-validation-for-a-covid-19-vaccine-and-emphasizes-need-for-equitable-global-access

9. Global Alliance for Vacccination and Immunization (GAVI). COVAX roll-out - Togo [Internet]. [cited 2021 Mar 24]. Available from: https://www.gavi.org/covax-vaccine-roll-out/togo

10. Staff S. Covid-19: India looking at all ‘serious’ side effects amid safety concerns about AstraZeneca vaccine [Internet]. Scroll.in. https://scroll.in; [cited 2021 Mar 26]. Available from: https://scroll.in/latest/989440/covid-19-india-looking-at-all-serious-side-effects-amid-safety-concerns-about-astrazeneca-vaccine

11. World Health Organization (WHO). WHO recommendation Serum Institute of India Pvt Ltd - COVID-19 Vaccine (ChAdOx1-S [recombinant]) - COVISHIELDTM [Internet]. WHO - Prequalification of Medical Products (IVDs, Medicines, Vaccines and Immunization Devices, Vector Control). 2021 [cited 2021 Mar 26]. Available from: https://extranet.who.int/pqweb/vaccines/covid-19-vaccine-chadox1-s-recombinant-covishield

12. COVID-19 national deployment and vaccination plan: Submission and review process, 29 January m2021 [Internet]. [cited 2021 Mar 30]. Available from: https://www.who.int/publications-detail-redirect/WHO-2019-nCoV-NDVP-country_plans-2021.1

13. République Togolaise, Ministère de la Santé, de l’Hygiène Publique et de l’Accès Universel aux Soins. Epidémie de COVID-19 au Togo-Rapport de situation N°381 au 28 mars 2021. 2021; 10p.

14. République Togolaise. COVID-19 [Internet]. COVID 19. [cited 2021 Mar 24]. Available from: https://covid19.gouv.tg/

15. République Togolaise. Ministère de santé, de l’hygiène publique et de l’accès universel aux soins. Vaccination du personnel de santé du pays contre la COVID-19 (Bulletin d’information N°2). 2021. 4p.

16. World Bank, Data Bank. CountryProfile [Internet]. [cited 2021 Mar 26]. Available from: https://databank.worldbank.org/views/reports/reportwidget.aspx?Report_Name=CountryProfile&Id=b450fd57&tbar=y&dd=y&inf=n&zm=n&country=TGO

17. République Togolaise, Direction Générale de la Statistique et de la Comptabilité Nationale du Togo (DGSCN). Résultats définitifs du 4ième Recensement Général de la Population et de l’Habitat (4ième RGPH4). Lomé : DGSCN; 2010, 66p. Disponible sur : https://inseed.tg/download/2958/.

18. Meo SA, Bukhari IA, Akram J, Meo AS, Klonoff DC. COVID-19 vaccines: comparison of biological, pharmacological characteristics and adverse effects of Pfizer/BioNTech and Moderna Vaccines. Eur Rev Med Pharmacol Sci. 2021 Feb;5(3):1663–9.

19. D’Alessandro D, Ciriminna S, Rossini A, Bossa MC, Fara GM. Requests of medical examinations after pneumococcal & influenza vaccination in the elderly. Indian J Med Res. 2004 May;119 Suppl:108–14.

20. Voysey M, Clemens SAC, Madhi SA, Weckx LY, Folegatti PM, Aley PK, et al. Safety and efficacy of the ChAdOx1 nCoV-19 vaccine (AZD1222) against SARS-CoV-2: an interim analysis of four randomised controlled trials in Brazil, South Africa, and the UK. Lancet Lond Engl. 2021;397(10269):99–111.

21. Torjesen I. Covid-19: First UK vaccine safety data are “reassuring,” says regulator. BMJ. 2021 Feb 8;372:363.

22. Ramasamy MN, Minassian AM, Ewer KJ, Flaxman AL, Folegatti PM, Owens DR, et al. Safety and immunogenicity of ChAdOx1 nCoV-19 vaccine administered in a prime-boost regimen in young and old adults (COV002): a single-blind, randomised, controlled, phase 2/3 trial. Lancet Lond Engl. 2021;396(10267):1979–93.

23. Ran L, Chen X, Wang Y, Wu W, Zhang L, Tan X. Risk Factors of Healthcare Workers With Coronavirus Disease 2019: A Retrospective Cohort Study in a Designated Hospital of Wuhan in China. Clin Infect Dis Off Publ Infect Dis Soc Am. 2020 Nov 19;71(16):2218–21.

24. Sadio AJ, Gbeasor-Komlanvi FA, Konu RY, Bakoubayi AW, Tchankoni MK, Bitty-Anderson AM, et al. Assessment of self-medication practices in the context of the COVID-19 outbreak in Togo. BMC Public Health. 2021 Jan 6;21(1):58.

25. Potluri T, Fink AL, Sylvia KE, Dhakal S, Vermillion MS, vom Steeg L, et al. Age-associated changes in the impact of sex steroids on influenza vaccine responses in males and females. Npj Vaccines. 2019 Jul 12;4(1):1–12.

26. Gee J. First Month of COVID-19 Vaccine Safety Monitoring — United States, December 14, 2020–January 13, 2021. MMWR Morb Mortal Wkly Rep [Internet]. 2021 [cited 2021 Mar 26];70. Available from: https://www.cdc.gov/mmwr/volumes/70/wr/mm7008e3.htm

27. Blumenthal KG, Robinson LB, Camargo CA, Shenoy ES, Banerji A, Landman AB, et al. Acute Allergic Reactions to mRNA COVID-19 Vaccines. JAMA [Internet]. 2021 Mar 8 [cited 2021 Mar 26]; Available from: https://jamanetwork.com/journals/jama/fullarticle/2777417

28. World Health Organization (WHO). Coronavirus disease (COVID-19): Vaccines safety [Internet]. [cited 2021 Mar 26]. Available from: https://www.who.int/news-room/q-a-detail/coronavirus-disease-(covid-19)-vaccines-safety

29. US Centers for Diseases Control and Prevention (US CDC). What to Expect after Getting a COVID-19 Vaccine [Internet]. Centers for Disease Control and Prevention. 2021 [cited 2021 Mar 26]. Available from: https://www.cdc.gov/coronavirus/2019-ncov/vaccines/expect/after.html

30. Government of the United Kingdom. Assests publishing service. COVID-19 vaccine AstraZeneca analysis print. 2021. [Cited 26/03/2021]. Available from: https://assets.publishing.service.gov.uk/government/uploads/system/uploads/attachment_data/file/972833/COVID-19_AstraZeneca_Vaccine_Analysis_Print.pdf.

31. World Health Organization (WHO). WHO statement on AstraZeneca COVID-19 vaccine safety signals [Internet]. [cited 2021 Mar 26]. Available from: https://www.who.int/news/item/17-03-2021-who-statement-on-astrazeneca-covid-19-vaccine-safety-signals

